# Interventions for the management of post COVID-19 condition (long COVID): Protocol for a living systematic review & network meta-analysis

**DOI:** 10.1101/2024.03.15.24304168

**Authors:** Dena Zeraatkar, Michael Ling, Sarah Kirsh, Tanvir Jassal, Tyler Pitre, Samantha Chakraborty, Tari Turner, Lyn Turkstra, Roger S. McIntyre, Ariel Izcovich, Lawrence Mbuagbaw, Thomas Agoritsas, Signe A. Flottorp, Paul Garner, Rachel Couban, Jason W. Busse

## Abstract

**Background:** Up to 15% of survivors of COVID-19 infection experience long-term health effects, including fatigue, myalgia, and impaired cognitive function, termed post COVID-19 condition or long COVID. Several trials that study the benefits and harms of various interventions to manage long COVID have been published and hundreds more are planned or are ongoing. Trustworthy systematic reviews that clarify the benefits and harms of interventions are critical to promote evidence-based practice.

**Objective:** To create and maintain a living systematic review and network meta-analysis addressing the benefits and harms of pharmacologic and non-pharmacologic interventions for the treatment and management of long COVID.

**Methods:** Eligible trials will randomize adults with long COVID, to pharmacologic or non-pharmacologic interventions, placebo, sham, or usual care. We will identify eligible studies by searches of MEDLINE, EMBASE, CINAHL, PsycInfo, AMED, and CENTRAL, from inception, without language restrictions.

Reviewers will work independently and in duplicate to screen search records, collect data from eligible trials, including trial and patient characteristics and outcomes of interest, and assess risk of bias. Our outcomes of interest will include fatigue, pain, post-exertional malaise, changes in education or employment status, cognitive function, mental health, dyspnea, quality of life, patient-reported physical function, recovery, and serious adverse events.

For each outcome, when possible, we will perform a frequentist random-effects network meta-analysis. When there are compelling reasons to suspect that certain interventions are only applicable or effective for a subtype of long COVID, we will perform separate network meta-analyses. The GRADE approach will guide our assessment of the certainty of evidence.

We will update our living review biannually, upon the publication of a seminal trial, or when new evidence emerges that may change clinical practice.

**Conclusion:** This living systematic review and network meta-analysis will provide comprehensive, trustworthy, and up-to-date summaries of the evidence addressing the benefits and harms of interventions for the treatment and management of long COVID. We will make our findings available publicly and work with guideline producing organizations to inform their recommendations.

## Background

The COVID-19 pandemic produced a global health crisis, affecting millions worldwide and causing significant health and economic consequences (1, 2). The prevalence of long COVID is difficult to establish, given most symptoms are nonspecific, and many studies lack sufficiently rigorous designs to confidently attribute symptoms to COVID-19 infection (3, 4). While most patients recover from COVID-19, evidence suggests that up to 15% of survivors may experience long-term health effects, including fatigue, myalgia, and impaired cognitive function—termed post COVID-19 condition or long COVID (5–14). The etiology of long COVID remains unclear and investigators have proposed several potential causes including viral persistence, autoimmunity, microclots, and psychological mechanisms (15).

There is heterogeneity in the definition of long COVID and some evidence indicates it may comprise several distinct phenotypes (16). Risk factors include sex, comorbidities, and patient-reported psychological distress (17–19). Conversely, severity of acute COVID-19 infection does not appear to predict long COVID and even non-hospitalized patients with mild infections are susceptible (20). Symptoms of long COVID may persist following acute infection or may relapse and remit (21). Evidence on the long term trajectory of the condition is limited but existing studies suggest that patients experience a significant reduction of symptoms at one year following the initial acute infection (22).

There is considerable overlap of signs, symptoms, and medical history between long COVID and myalgic encephalomyelitis/chronic fatigue syndrome (ME/CFS) (23). For example, ME/CFS often emerges following viral infections, similar to long COVID that emerges following infection with SARS-CoV-2 (23). Notably, approximately half of patients diagnosed with long COVID also meet criteria for ME/CFS (24–26).

Considerable resources have been invested to study long COVID, including $1 billion by the US National Institutes of Health (NIH) (27). In August 2023, the NIH established the Office of Long COVID Research and Practice and launched a suite of clinical trials investigating treatments, among them five adaptive platform trials—trials that compare several interventions simultaneously with the option to add or remove interventions based on emerging evidence (28, 29).

Several trials testing interventions for the management of long COVID have been published to date (30–33) and hundreds more are planned or ongoing (34–36). These trials, however, will be published faster than evidence users, such as clinicians and patients, can read them or make sense of them and will come with strengths and limitations that may not be immediately apparent.

Ongoing discourse in the literature shows that there is uncertainty about optimal treatment modalities for long COVID (37–39). In the absence of trustworthy summaries of the evidence, patients living with long COVID are receiving unproven treatments—many of which are costly and some of which may be harmful (37–41). For interventions for which trials have been published, for example trials testing exercise and cognitive behavioral therapy, patients and healthcare providers have questioned their credibility (42–44). Trustworthy systematic reviews that clarify the benefits and harms of available interventions are critical to promote the evidence-based care of patients.

We present a protocol for a living systematic review and network meta-analysis of all pharmacologic and non-pharmacologic interventions for long COVID. Unlike a traditional systematic review that is only up-to-date at a single point in time, we will update this living review as new evidence emerges (45, 46). This review will provide comprehensive, trustworthy, and up-to-date summaries of the evidence addressing interventions for the management of long COVID.

We anticipate that the living systematic review and network meta-analysis will become a trusted reference point for clinicians, patients, and national and international professional associations and authoritative organizations that intend to produce guideline recommendations on the treatment and management of long COVID. We have already engaged a committee of evidence users, including healthcare professionals and guideline-producing organizations, in the design of this initiative. This committee will frequently be asked for feedback on our methods and the presentation of our results, to ensure that our products align with their needs. We hope that our findings will expedite the identification of the most effective interventions for patients with long COVID and inform future guideline development efforts.

## Methods

We report our protocol according to the Preferred Reporting Items for Systematic Reviews and Meta-Analyses (PRISMA) extension for protocols (47, 48).

### Eligibility criteria

Eligible studies will randomize adults with long COVID, defined by the World Health Organization (WHO) as symptoms at three or more months following laboratory confirmed, probable, or suspected COVID-19 infection and that persist for at least two months, to pharmacologic or non-pharmacologic interventions, placebo, sham, usual care, or to alternative pharmacologic or non-pharmacologic interventions, without any restrictions on date or language of publication (21). Given the diverse manifestations of long COVID, our review will not further restrict eligibility based on diagnostic criteria (21). Subgroup analyses will, however, investigate the differences in the effectiveness of interventions according to diagnostic criteria.

We anticipate that trials will likely largely include patients that meet the WHO criteria for long COVID but some trials may not report the time since the acute COVID-19 infection or duration of long COVID symptoms. We will include such trials in our primary analysis but will also perform additional sensitivity analyses that exclude these trials to test the robustness of our findings.

Based on empirical evidence that preprint and published reports of randomized trials generally provide consistent results (49–51), we will include both preprint and published trial reports, but will also perform sensitivity analyses excluding preprints (49, 52).

At this time, there is too little known about long COVID to anticipate which interventions may be effective. Trials are investigating many types of interventions, including drugs (e.g., colchicine, sulodexide, beta-blockers), behavioral and physical therapies (e.g., cognitive behavioral therapy, aerobic exercise training), dietary supplements, and medical devices (transcranial direct current stimulation) (28, 35, 53, 54). We will not restrict eligibility based on the type of intervention and anticipate including trials addressing a diverse range of therapies.

We will exclude pseudorandomized trials, trials involving animals, trials investigating treatments for acute COVID-19, trials testing interventions to prevent long COVID, and trials including patients that do not meet criteria for long COVID (21, 55). We will also exclude randomized trials with fewer than 25 participants in each arm. We anticipate that smaller trials are unlikely to meaningfully contribute to meta-analyses, are more likely to include unrepresentative samples, arms that are prognostically imbalanced, and are at higher risk of publication bias (56).

While there are hundreds of long COVID trials underway, trials have progressed at a slower pace than anticipated (57). Depending on feasibility, for select interventions of high potential interest to evidence users, in the first year of the living systematic review, we will consider including indirect evidence from trials addressing interventions for ME/CFS. The decision to consider indirect evidence from ME/CFS will be made with input from clinical experts and by evidence users. Subgroup analyses will investigate differences between the effects of interventions for long COVID and ME/CFS. If we find evidence that the effects of interventions are inconsistent between patients with long COVID and ME/CFS, we will only present results of trials addressing long COVID. Similar to long COVID, we will include trials that recruit patients according to published diagnostic criteria for ME/CFS but perform subgroup analyses investigating potential differences in the effects of interventions based on these diagnostic criteria.

### Data sources and search strategy

An experienced medical research librarian developed a comprehensive search strategy for MEDLINE, EMBASE, CINAHL, PsycInfo, AMED, CENTRAL, and preprint servers (MedRxiv and ResearchSquare), from inception (Appendix). Our search strategy combines terms related to long COVID with a filter for randomized trials. Additionally, we plan to supplement our search using the Epistemonikos COVID-19 Repository—a living catalogue of COVID-19 research—and by reviewing the references of relevant systematic reviews (32, 35, 58).

The medical research librarian will update the searches at minimum biannually to facilitate biannual updates of the living review. We aim to update the search no more than three months before the publication of each iteration of the review.

To ensure our review remains up-to-date, for select interventions identified by our network of evidence users as highly important with great potential for efficacy, we will integrate methods for prospective systematic reviews—systematic reviews that include unpublished and interim data from ongoing and completed trials (59). During the COVID-19 pandemic, trialists reported trouble publishing trials due to null results, inability to achieve the target sample size, and waning interest from journals. In response, the WHO REACT Working Group performed prospective systematic reviews for select topics (60, 61). These reviews sourced ongoing trials via trial registries and invited trialists to share their interim or completed data. This model proved successful and also mitigated the influence of publication bias on review results (60, 61). We plan to emulate a similar model for critical interventions of long COVID. Our experience with other living systematic reviews and the experience of other research groups suggest that some trial groups are receptive to sharing unpublished trial data (60–62).

For interventions for which we consider evidence from ME/CFS trials, we will work with an experienced research medical librarian to devise additional search strategies specific to the interventions for which we will consider indirect evidence.

### Study selection

Following training and calibration exercises to ensure sufficient agreement, pairs of reviewers will work independently and in duplicate to screen the titles and abstracts of search records and subsequently the full-texts of records deemed eligible at the title and abstracts screening stage using Covidence (https://www.covidence.org), an online systematic review software for screening titles and abstracts and full-text articles. Reviewers will resolve disagreements by discussion, or, if necessary, by adjudication by a third reviewer.

### Data extraction

Following training and calibration exercises to ensure sufficient agreement, pairs of reviewers will work independently and in duplicate to collect data from eligible trials using a pilot-tested data collection form in an Excel spreadsheet (Microsoft Office Excel 2019). Reviewers will resolve disagreements by discussion or by consultation with a third reviewer, if necessary.

Reviewers will collect data on trial characteristics (e.g., trial design, country of origin, funding, diagnostic criteria for long COVID), patient characteristics (e.g., age, sex and gender, employment and education status, receipt of COVID-19 vaccination, method of acute COVID-19 diagnosis, severity of acute COVID-19 infection, duration of long COVID, number of infections, equity-related characteristics, long COVID symptoms), characteristics of interventions and comparators (e.g., type of intervention, treatment duration), and patient-important outcomes.

For dichotomous outcomes, reviewers will extract the number of patients and events in each arm, and, for continuous outcomes, the number of patients, a measure of central tendency (mean or median), and a measure of variability (e.g., standard deviation, standard error, 95% confidence interval, p-value) for each arm. For continuous outcomes, reviewers will prioritize extracting changes in the outcome from baseline, and if not reported, the outcome at follow-up. For each outcome, reviewers will preferentially extract the results from intention-to-treat analyses without any imputations for missing data, when reported.

We will also prioritize outcome data at the longest reported timepoint at which the intervention is still being administered to allow for potential cumulative effects of interventions to emerge without effects waning due to termination of the intervention. When trials report data at timepoints at which the intervention is no longer being administered but randomized groups are still maintained, we may consult experts who are blind to the results of the trial about the duration of time the interventions being tested are expected to exert effects. If appropriate, we will consider extracting and analyzing data at the longest reported point of follow-up even if the intervention is no longer administered.

Our outcomes of interest are informed by a published core outcome set for long COVID and will include at minimum fatigue, pain, post-exertional malaise, changes in education or employment status, cognitive function, mental health, dyspnea, quality of life, patient-reported physical function, recovery, and serious adverse events (as defined by each trial) (63, 64). We will extract data for all validated instruments measuring our outcomes of interest. Table 1 presents examples. We will re-evaluate and potentially revise our choice of outcomes annually by discussion with patient partners and Knowledge Users and by considering emerging evidence. If we find important reasons to include additional outcomes, we will revisit previously extracted trials to collect data on additional outcomes.

**Table 1:**
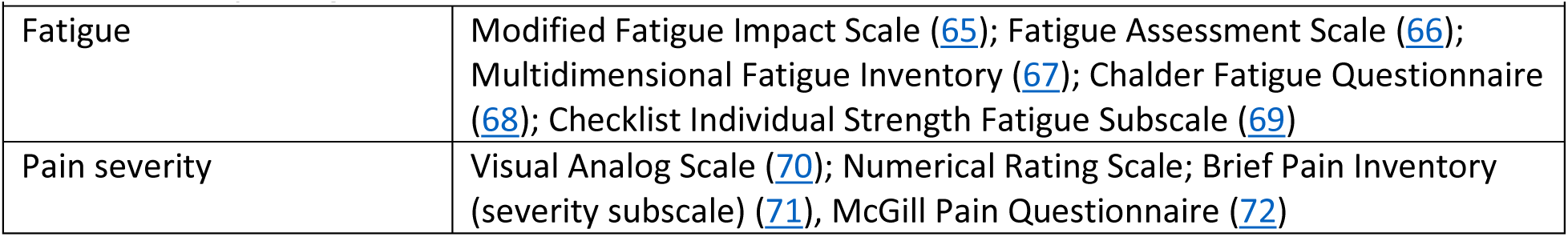
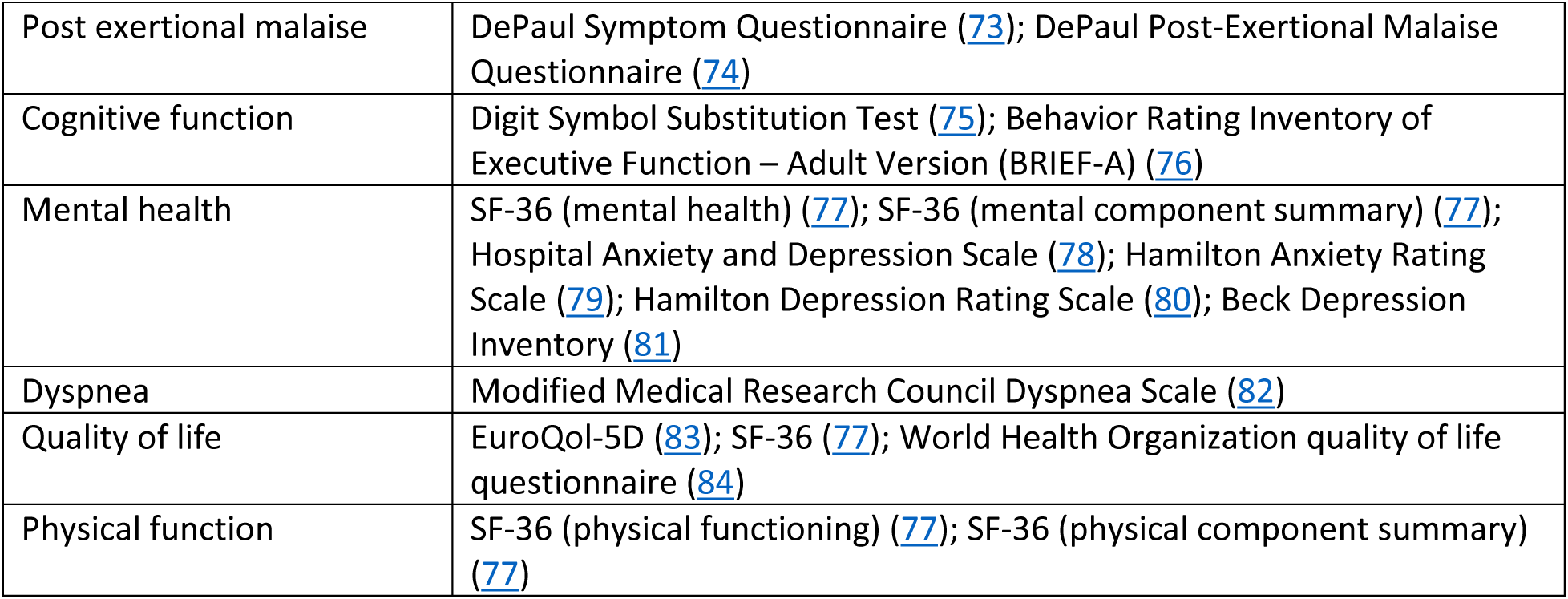
Example eligible instruments for outcomes of interest.

| Table 1: Example eligible instruments for outcomes of interest |  |
| --- | --- |
| Fatigue | Modified Fatigue Impact Scale (65); Fatigue Assessment Scale (66); Multidimensional Fatigue Inventory (67); Chalder Fatigue Questionnaire (68); Checklist Individual Strength Fatigue Subscale (69) |
| Pain severity | Visual Analog Scale (70); Numerical Rating Scale; Brief Pain Inventory (severity subscale) (71), McGill Pain Questionnaire (72) |
| Post exertional malaise | DePaul Symptom Questionnaire (73); DePaul Post-Exertional Malaise Questionnaire (74) |
| Cognitive function | Digit Symbol Substitution Test (75); Behavior Rating Inventory of Executive Function – Adult Version (BRIEF-A) (76) |
| Mental health | SF-36 (mental health) (77); SF-36 (mental component summary) (77); Hospital Anxiety and Depression Scale (78); Hamilton Anxiety Rating Scale (79); Hamilton Depression Rating Scale (80); Beck Depression Inventory (81) |
| Dyspnea | Modified Medical Research Council Dyspnea Scale (82) |
| Quality of life | EuroQol-5D (83); SF-36 (77); World Health Organization quality of life questionnaire (84) |
| Physical function | SF-36 (physical functioning) (77); SF-36 (physical component summary) (77) |

Given the relapse and remission patterns associated with long COVID and the potential for interventions to have long term effects, for crossover trials, we will only collect data for the first phase of the trial before washout and crossover of patients.

For trials initially available as a preprint that are subsequently published, we will rely on the results of the published trial unless the preprint reports additional outcome data not included in the published trial report.

In response to growing concerns about the prevalence of potentially fabricated or falsified research (85, 86), reviewers will use the TRACT checklist to evaluate each trial for the risk of data falsification or fabrication (87). This checklist includes 19 items in seven domains (governance, author group, plausibility of intervention, timeframe, dropouts, baseline characteristics, and outcomes) addressing the integrity and trustworthiness of trials. The checklist does not include a cutoff at which a trial is considered suspicious and there is currently limited experience using the checklist in systematic reviews. Therefore, the authorship group will discuss all trials that are flagged as raising concerns for one or more domains. We will perform sensitivity analyses excluding trials that are deemed suspicious. We are also aware of another instrument to assess the risk of falsified or fabricated data in trials currently under development (88). Upon its publication, we will review the instrument and consider incorporating in our workflow.

We anticipate that the effects of interventions may depend on diagnostic criteria for long COVID or ME/CFS, severity of acute COVID-19, time since infection, number of infections, vaccination status, and SARS-CoV-2 variant (17). When reported, we will extract stratified data based on these factors to facilitate subgroup analyses.

### Risk of bias assessment

Following training and calibration, reviewers will work independently and in duplicate to assess risk of bias using a modified version of the Cochrane-endorsed Risk of Bias (RoB) 2.0 tool—the gold standard for assessing limitations in trials that may bias results (89). The RoB 2.0 assesses the risk of bias across five domains: bias due to randomization (e.g., random sequence generation, allocation concealment), bias due to deviations from the intended intervention (e.g., lack of blinding leading to imbalances in co-interventions across trial arms), bias due to missing outcome data (e.g., attrition), bias due to measurement of the outcome (e.g., unblinded outcome assessment), and selective reporting (e.g., selective reporting of outcome measures based on results).

To assess the risk of bias due to deviations from the intended intervention, we will consider the effect of assignment rather than adherence to the intervention—since this effect is likely to be the observed effect in clinical settings and of the greatest interest to evidence users. Our modified version of the tool includes the same domains, but with revised response options (i.e., definitely low risk of bias, probably low risk of bias, probably high risk of bias, and definitely high risk of bias) and guidance tailored to issues relevant for the present review (e.g., removing guidance for assessing risk of bias of adhering to the intervention, listing important cointerventions).

Unless we encounter compelling reasons to do otherwise, we will consider trials without blinding of patients, healthcare providers, and investigators at high risk of bias due to deviations from intended intervention. This decision is based on the potential for unequal distribution of potentially effective cointerventions (e.g., physical activity, social engagement, energy management) across trial arms.

Reviewers will resolve disagreements by discussion or by consultation with a third reviewer, if necessary.

### Data synthesis and analysis

To describe trials and participants, we will present descriptive characteristics. Means and medians and associated measures of variability (e.g., 95% CI, SD) will describe continuous variables and counts and proportions dichotomous and categorical variables.

Given the heterogeneity in the definition of long COVID and evidence indicating that it may comprise several distinct phenotypes (16), we anticipate that some interventions may be better suited for patients with certain phenotypes of long COVID. For example, pulmonary rehabilitation will likely only be effective for patients who are experiencing pulmonary sequelae related to COVID-19, (90), interventions targeting cognitive function will likely only be effective for patients experiencing neurocognitive sequelae related to COVID-19 (91), and interventions aimed at restoring sense of smell will only be effective for patients experiencing anosmia (92). Other interventions may be suitable for patients experiencing general symptoms related to long COVID, such as fatigue, post-exertional malaise, and headaches. We will perform separate syntheses when there are compelling clinical reasons to suspect that certain interventions are only applicable or effective for a subtype of long COVID. Clinical experts in our authorship group will lead these decisions and regularly revisit them based on new evidence.

To summarize the comparative efficacy and harms of interventions, we will perform network and pairwise meta-analyses. Network meta-analyses compare three or more interventions, grouped into nodes, by pooling direct and indirect evidence (59). To facilitate network meta-analysis, we will classify interventions into “nodes” considering the drug class for pharmacologic interventions, class of therapy (e.g., cognitive behavioral therapy, aerobic exercise) for behavioral and physical interventions, and characteristics of the therapy for other non-pharmacologic interventions. We will group all therapeutic doses of the same drug class in the same node. If we find evidence that the effects of interventions are different based on their mode (group versus individual, online or in-person), intensity, or dose of delivery, we will create separate nodes. We will group placebo and sham interventions and standard care in the same node (93).

We anticipate that trials may measure the same constructs using different instruments. We plan to pool the results of each unique instrument separately. We will avoid converting effects across instruments, due to potential differences in the range of the construct covered by each instrument, or using standardized mean difference, due to its potential to be influenced by differences in variability across trial populations (94).

For each outcome and outcome measure, random-effects network meta-analysis using a frequentist framework with the restricted maximum likelihood (REML) heterogeneity estimator—a conservative approach to network meta-analysis—will pool the results of trials (95, 96). Our choice of a frequentist over a Bayesian framework is motivated by evidence that there are seldom important differences between the results of Bayesian and frequentist networks and the computational complexity associated with analyzing large networks under a Bayesian framework (95).

Relative risks (RRs) will summarize the results of dichotomous outcomes—except for serious adverse events that will be summarized using risk difference (RD) due to the propensity for trials to frequently report 0 events which precludes calculation of RRs and odds ratios—and mean differences (MDs) continuous outcomes, along with associated 95% confidence intervals. When network meta-analysis is not possible, we will perform pairwise random-effects meta-analysis with the REML heterogeneity estimator (96).

We will use I^2^ statistics to summarize the magnitude of heterogeneity in meta-analyses and interpret an I^2^ value of 0% to 40% as not important, 30% to 60% as moderate heterogeneity, and 50% to 90% as substantial heterogeneity, and 75% to 100% as considerable heterogeneity (97, 98). The I^2^ value, however, is prone to misinterpretation since even small degrees of unimportant inconsistency may translate to high I^2^ values if estimates from studies are highly precise (99, 100). Hence, we will also consider the absolute magnitude of differences in effect estimates across studies. For analyses that include 10 or more studies, we will test for publication bias using Egger’s test and qualitatively review funnel plots for evidence of asymmetry (101, 102).

A feature of network meta-analyses is their ability to generate treatment rankings. However, based on empirical evidence and theoretical considerations that suggest that ranking methods for network meta-analyses may be misleading, we will avoid interpreting our results based on rankings (103–105).

Diverse trial outcomes or outcome measures may lead to disconnected networks or preclude network formation. In such situations, consistent with established guidance, we will present the results of disconnected networks separately (59). We refrain from model-based approaches to link networks because of their novelty and limited evidence supporting their reliability (106). If networks cannot be formed, we will present pairwise meta-analyses. With the emergence of more trial evidence, we anticipate that disconnected networks will eventually combine to become connected.

To enhance interpretability of results, we will transform relative effects (e.g., RRs) to absolute effects (e.g., number of events per 1,000 patients), using the median risk of the outcome reported across the control groups of trials (107).

We will test for incoherence using node-splitting (108). In case of incoherence, we will investigate potential sources considering our *a priori* defined hypotheses for effect modification: diagnostic criteria for long COVID, time since infection, number of infections, vaccination status, severity of acute COVID-19, and SARS-CoV-2 variant (17). Should we choose to include evidence from ME/CFS trials, we will also consider ME/CFS as a potential source of incoherence. In the event that tests for effect modification are unable to identify a credible source of incoherence, we will downgrade the certainty of evidence. Conversely, if we confidently identify the source of incoherence, we will perform separate analyses based on that source.

We have also generated similar *a priori* factors to explain potential heterogeneity in results across trials: diagnostic criteria for long COVID, time since infection, number of infections, vaccination status, severity of acute COVID-19, and SARS-CoV-2 variant (17, 109). Meta-regressions and subgroup analyses will test for subgroup effects based on these factors. Notably, we will avoid pooling trials rated at low and high risk of bias indiscriminately. Instead, we will test for differences between the results of these trials, and when we detect important differences, rely on trials at low risk of bias. If we choose to include evidence from ME/CFS trials, we will also perform subgroup analyses comparing the results of trials addressing long COVID and ME/CFS.

Inferences of subgroup effects often prove spurious (110–119). Such spurious claims may be attributed to testing many factors, leading to apparent but inaccurate evidence of effect modification due to chance, selective reporting, or improper statistical analysis (119–125). To avoid misleading claims, we will assess the credibility of subgroup effects using the ICEMAN tool—the gold standard tool for evaluating effect modification in trials and systematic reviews (126).

We will perform all analyses using the *meta* and *netmeta* packages in R (Vienna, Austria; Version 4.1.2) and make all code to reproduce our results freely accessible on Open Science Framework (127, 128).

### Certainty of evidence and reporting

Results from studies may appear impressive, but we may not have much confidence in those results due to the limitations of the evidence. The GRADE approach for network meta-analysis will guide our assessment of the certainty (quality) of evidence (129–131). The GRADE approach rates the evidence as either high, moderate, low, or very low certainty based on considerations of risk of bias (i.e., study limitations), inconsistency (i.e., heterogeneity in results across trials), indirectness (i.e., differences between the questions addressed in studies and the question of interest), publication bias (i.e., the tendency for studies with statistically significant results or positive results to be published, published faster, or published in journals with higher visibility), imprecision (i.e., random error), intransitivity (i.e., violation of joint randomizability), and incoherence (i.e., differences between direct and indirect estimate). High certainty evidence indicates situations in which we are confident that the estimated effect represents the true effect and low or very low certainty evidence indicates situations in which the estimated effect may be substantially different from the true effect.

A minimally contextualized approach will guide our judgments related to imprecision (132). The minimally contextualized approach does not consider statistical significance as the only indicator of whether an intervention is effective. An estimate may not be statistically significant but may still have evidence of moderate certainty for benefit or harm, depending on whether the confidence intervals cross the thresholds of clinical significance. Conversely, an intervention may produce results that are statistically significant but that indicate no important benefit or harm. The minimally contextualized approach considers only whether the effect estimates exceed the minimum important difference (MID)—the smallest difference in an outcome that patients find important—and does not consider whether the effect is small, moderate, or large.

We will source MIDs either from the literature, or, when not available, survey our review authors and patient partners. MIDs of patient-reported outcomes are typically determined either using anchor-based methods or distribution-based methods (133). Anchor-based methods use an external "anchor" to interpret the magnitude of change in a measure or outcome. Distribution-based methods rely on the distribution of the data to interpret the importance of change in a measure. We will prioritize anchor-based MIDs over distribution-based MIDs, since anchor-based estimates reflect patients’ direct experiences. We anticipate that guideline producing organizations will fully contextualize the results to formulate recommendations (134). Finally, should a published MID be unavailable for any of the outcomes of interest, particularly for dichotomous outcomes like recovery and return to work/education for which MIDs are typically not derived, we will survey patient partners and our partner evidence users on reasonable ranges using a previously established process (135).

To make judgements about intransitivity, we will consider the potential effect modifiers within the network, including the credibility of effect modification, the strength of effect modification, and the distribution of effect modifiers across direct comparisons (136). There is currently limited evidence on potential effect modifiers of interventions for long COVID. We anticipate that the effects of interventions may vary based on diagnostic criteria, severity of acute COVID-19, time since infection, number of infections, vaccination status, and SARS-CoV-2 variant. If we find credible evidence of effect modification based on these or other factors, we will consider rating down for intransitivity when appropriate. For comparisons that include evidence from ME/CFS trials, we will additionally rate down for indirectness.

### Reporting of results

We will report our living review according to the PRISMA checklist for network meta-analyses (47, 48). PRISMA flow diagrams will illustrate the total number of search records, the number of records excluded, reasons for exclusion, and the total number of trials included in the review. Network and forest plots will present network geometry and results from meta-analyses, respectively. GRADE Evidence Profiles will summarize effect estimates and the certainty of evidence (107).

We will describe our results using GRADE plain language summaries (i.e., describing high certainty evidence with declarative statements, moderate certainty evidence with ‘probably’, low certainty evidence with ‘may’ and very low with ‘very uncertain’) (137).

For each outcome, we will place interventions in categories from best to worst (138, 139). First, we will classify interventions according to whether they are more or less effective than placebo or standard care. If the 95% confidence intervals of effect estimates comparing interventions with placebo or standard care cross the MID, the intervention will remain in the same group as placebo or standard care. If, on the other hand, the interval does not cross the MID, the intervention can be classified as more or less effective than placebo or standard care, depending on the direction of the effect. Subsequently, we will compare the interventions classified as more effective than placebo or standard care against each other by examining whether the differences in effects between them exceed the MID. In the final step, we will further categorize interventions according to their certainty of evidence.

Each iteration of the living review will also be accompanied by a plain language summary (each <800 words) that describe our findings in simple language for healthcare providers and patients who may not be familiar with network meta-analysis methods. We will draft these summaries according to established guidance by Cochrane: they will describe the types of interventions tested and describe the benefits and harms of interventions supported by moderate or high certainty evidence (59). We anticipate that these plain language summaries will reduce the opportunity for misinterpretation of our findings by healthcare providers, patients, and other decision-makers who may not be familiar with network meta-analysis methods or the interpretation of evidence according to the GRADE approach. For each iteration of the review, we will engage three to five patient partners and three to five healthcare providers to review the plain language summaries for readability and acceptability.

### Updating and triggers for retirement

We will update our living systematic review and network meta-analysis every 6 months or sooner in the event of the publication of practice-changing evidence (e.g., publication of a seminal trial, when the certainty of evidence or the magnitude or direction of the effect of an intervention importantly changes). Preprints and journal publications will communicate the results of each iteration of our review. This approach adheres to best practices in updating living reviews: it balances the need for up-to-date evidence with the time needed to ensure that the review is sufficiently rigorous, focuses our efforts on disseminating critical findings, and maximizes the feasibility of the project (45, 140).

We will retire the living systematic review when the evidence base becomes stable with few to no new trials, if we reach high or moderate certainty evidence for all interventions and outcomes suggesting that new evidence is unlikely to change current estimates, when our network of evidence users suggest that the findings of the living systematic review are no longer relevant to them, or if we deplete our funding, or can no longer maintain the personnel needed to continue the living review (141). At this time, given the timeline of planned trials and the funding available to us, we intend to continue to maintain the living systematic review for three years (28, 29).

Our Open Science Framework repository dedicated to the living review will inform evidence users of the status of the review (whether it is active or retired), the anticipated date of the next update, and the results of the most recent iteration of the review (osf.io/9h7zm).

### Patient involvement

As part of our funding application, the Long Covid Web Patient Advisory Council reviewed and offered feedback on the protocol.

When interpreting our results, we will rely on patients’ judgements about whether they consider benefits of a particular therapy to outweigh harms. To do this, we will perform semi-structured interviews with purposively selected groups of patient partners, aiming for diversity in demographics (e.g., age, sex, underrepresented racial or ethnic groups, income, abilities) and experiences of long COVID (e.g., severity, duration). These interviews will be intended to offer explanations for why certain therapies may be preferential to others. Consistent with guidance for qualitative research, we will involve enough patient partners until we reach saturation and no new insights are provided (143). Our previous experiences performing similar semi-structured interviews suggest that we will need to involve at minimum six to seven patients for each iteration of the review.

For each iteration of the review, three to four patient partners will also review plain language summaries that describe our findings using language that will be accessible to the general public for readability and acceptability (144, 145).

We anticipate that the living systematic review will become a trusted reference point for national and international professional associations and authoritative organizations that intend to produce guideline recommendations on the management of long COVIDs. We will prioritize engaging organizations that involve patients in the guideline development process and consider patient values and preferences— consistent with standards for producing trustworthy guidelines (146–149).

## Discussion

### Anticipated findings

Our living systematic review and network meta-analysis will provide comprehensive, trustworthy, and up-to-date summaries of the evidence addressing interventions for the management of long COVID. We expect to produce at minimum six iterations of the living review. Each iteration will be accompanied by plain language summaries for patients, healthcare providers, and other decision-makers who may not be familiar with network meta-analysis methods. We hope that our findings will accelerate the identification of the most effective interventions for patients with long COVID.

To our knowledge, this is the first living network meta-analysis investigating the benefits and harms of pharmacologic and non-pharmacologic interventions for long COVID. Discussions with our network and searches of research databases confirm that the proposed review is original and there are no existing reviews of the same scope or rigor as we propose. The Canadian Agency for Drugs and Technologies in Health (CADTH) living review only provides a descriptive summary of trials, without quantitative synthesis or rigorous appraisal (34). Other living reviews addressing long COVID do not focus on interventions for management of the condition, perform network meta-analysis, or evaluate the certainty of evidence (150).

We anticipate that this living systematic review and network meta-analysis will become a trusted reference point for clinicians, patients, and national and international professional associations and authoritative organizations that intend to produce guideline recommendations on the treatment and management of long COVIDs. We invite guideline producing organizations that are either publishing or planning clinical practice guidelines addressing long COVID to join our committee of evidence users, who inform the type of data that we collect and our methodological approaches to ensure that our products align with their needs. Our model of simultaneously providing trustworthy summaries of evidence to several guideline developing organizations will prove more efficient than each organization performing its own overlapping evidence syntheses and optimize the translation of research into clinical practice.

We also invite clinical trialists to share interim or completed trial data for inclusion in the living systematic review. We are especially interested in trials that may not be published due to null findings, insufficient sample sizes, or lack of interest from journals—trials at risk of the "file drawer" effect. Sharing this data helps prevent publication bias and honors the commitment of trial participants, funders, and investigators by ensuring that their efforts improve patient care.

### Strengths and limitations

Strengths of this living systematic review and network meta-analysis include a broad search strategy and inclusion criteria, consideration of outcomes of interest reflecting the values and preferences of patients, screening of studies and extraction of data in duplicate to reduce the opportunity for errors (151–156), application of GRADE to evaluate the certainty of evidence, and commitment to data sharing and open science practices (157).

We also acknowledge potential limitations. First, despite our comprehensive search strategy, it is possible we will not identify all eligible randomized trials. To mitigate this issue, we will supplement our search with the Epistemonikos COVID-19 repository, which independently performs searches and screens for relevant randomized trials (58).

Second, while there are many trials underway, trials have progressed at a slower pace than anticipated (57). Our ongoing surveillance of trial registries has identified over 200 eligible trials, indicating that considerable trial evidence is forthcoming. For example, the US RECOVER program includes five adaptive platform trials (158). In 2023, Canada funded a $20 million research network, called Long Covid Web, which is also anticipated to support clinical trials (159). Further, RECLAIM, the Canadian platform trial investigating interventions for long COVID, will also contribute evidence for several therapeutic interventions. Our plan to integrate methods for prospective systematic reviews will also ensure that there will be sufficient data to make a living review both feasible and valuable for evidence users.

Finally, we have made certain methodological decisions to ensure the feasibility of our living systematic review. One such decision is our approach to evaluating the risk of bias due to missing outcome data.We plan to assess the risk of bias due to missing outcome data by considering the proportion of participants with missing outcome data, reasons for missingness, and whether missing data could importantly influence the effect estimate. An alternative approach to assessing risk of bias due to missing outcome data involves imputing missing data across a range of plausible scenarios and making a judgment based on the robustness of the results to imputation (160). This approach, however, is also impractical due to the anticipated numbers of outcomes and comparisons in the living systematic review and network meta-analysis (160). We believe the proposed methods strike a reasonable balance between rigor and feasibility.

## Conclusion

This protocol describes the planned methods of a living systematic review and network meta-analysis addressing the comparative effectiveness of interventions for the management of long COVID. We anticipate that our findings will inform clinical practice, clinical practice guidelines and guide the investigation of promising interventions for future trials.

## Data Availability

Data: Not applicable.

https://osf.io/9h7zm/

