## Supplement for "Interventions for the management of post COVID-19 condition (long COVID): Protocol for a living systematic review & network meta-analysis"

Summary of search and strategies long COVID RCTs

| MEDLINE | 3145 |
| --- | --- |
| Embase | 3855 |
| PsycInfo | 96 |
| AMED | 14 |
| CINAHL | 733 |
| Central | 6088 |
| Subtotal | 13931 |
| -dupes | -2257 |
| Total | 11674 |

Search strategy adapted from Campbell SM. Filter to Retrieve Studies Related to Long COVID in the OVID Medline Database. John W. Scott Health Sciences Library, University of Alberta, Rev. Jan 10, 2023.. [https://docs.google.com/document/d/1iEN3WvRGAtj_NF60LUgElBA4FEcCdOgW1gcadK45ltE/edit#](https://docs.google.com/document/d/1iEN3WvRGAtj_NF60LUgElBA4FEcCdOgW1gcadK45ltE/edit)

Nov 30, 2023

MEDLINE (OVID)

Database: OVID Medline Epub Ahead of Print, In-Process & Other Non-Indexed Citations, Ovid MEDLINE(R) Daily and Ovid MEDLINE(R) 1946 to Present

Search Strategy:

--------------------------------------------------------------------------------

1	Post-Acute COVID-19 Syndrome/ (2693)

2	(long* adj3 (covid or covid-19 or covid19 or sars cov 2)).mp. [mp=title, book title, abstract, original title, name of substance word, subject heading word, floating sub-heading word, keyword heading word, organism supplementary concept word, protocol supplementary concept word, rare disease supplementary concept word, unique identifier, synonyms, population supplementary concept word, anatomy supplementary concept word] (6413)

3	((covid* or "corona virus 2019" or "coronavirus2019" or SARS-CoV-2 or sars cov 2) adj3 (syndrome or persist* or lingering or chronic or ongoing or long-term or "long term" or long-haul or "long haul" or convalescen* or rehabilitat*)).ti. (5492)

4	((exp SARS-CoV-2/ or exp COVID-19/) and sequela*.ti,ab.) or ("long Covid" or ((Covid or Covid19 or "corona virus 2019" or "coronavirus 2019" or SARS-CoV-2 or "B.1.1.7" or "B.1.351" or "B.1.1.28" or "B.1.617" or "BA.1" or "BA.2" or "BA.3" or "BA.4" or "BA.5" or omicron or deltacron or "delta variant" or "delta subvariant" or "XBB.1.3") adj3 (PASC or sequela* or "post acute" or postacute or prolonged or "long haul*" or chronic or lingering or ongoing or persistent or "long term" or "more than 12 weeks" or "more than 24 weeks"))).mp. (14366)

Annotation: Sandra Campbell filter

5	1 or 2 or 3 or 4 (18940)

6	randomized controlled trial.pt. (604012)

7	controlled clinical trial.pt. (95469)

8	randomi?ed.ab. (747389)

9	placebo.ab. (243520)

10	drug therapy.fs. (2643985)

11	randomly.ab. (421817)

12	trial.ab. (674743)

13	groups.ab. (2602907)

14	or/6-13 (5837395)

15	exp animals/ not humans.sh. (5174725)

16	14 not 15 (5102329)

17	5 and 16 (3145)

Embase (OVID)

Database: Embase <1974 to 2023 November 29>

Search Strategy:

--------------------------------------------------------------------------------

1	long COVID/ (6134)

2	(long* adj3 (covid or covid-19 or covid19 or sars cov 2)).mp. [mp=title, abstract, heading word, drug trade name, original title, device manufacturer, drug manufacturer, device trade name, keyword heading word, floating subheading word, candidate term word] (9976)

3	((covid* or "corona virus 2019" or "coronavirus2019" or SARS-CoV-2 or sars cov 2) adj3 (syndrome or persist* or lingering or chronic or ongoing or long-term or "long term" or long-haul or "long haul" or convalescen* or rehabilitat*)).ti. (6624)

4	exp coronavirus disease 2019/ and sequela*.ti,ab. (4716)

5	((Covid or Covid19 or "corona virus 2019" or "coronavirus 2019" or SARS-CoV-2 or "B.1.1.7" or "B.1.351" or "B.1.1.28" or "B.1.617" or "BA.1" or "BA.2" or "BA.3" or "BA.4" or "BA.5" or omicron or deltacron or "delta variant" or "delta subvariant" or "XBB.1.3") adj3 (PASC or sequela* or "post acute" or postacute or prolonged or "long haul*" or chronic or lingering or ongoing or persistent or "long term" or "more than 12 weeks" or "more than 24 weeks")).mp. [mp=title, abstract, heading word, drug trade name, original title, device manufacturer, drug manufacturer, device trade name, keyword heading word, floating subheading word, candidate term word] (12484)

6	or/1-5 (24627)

7	randomized controlled trial/ (795458)

8	Controlled clinical study/ (471569)

9	random$.ti,ab. (2003505)

10	randomization/ (98950)

11	intermethod comparison/ (302807)

12	placebo.ti,ab. (369125)

13	(compare or compared or comparison).ti. (611285)

14	((evaluated or evaluate or evaluating or assessed or assess) and (compare or compared or comparing or comparison)).ab. (2822956)

15	(open adj label).ti,ab. (111156)

16	((double or single or doubly or singly) adj (blind or blinded or blindly)).ti,ab. (276603)

17	double blind procedure/ (213168)

18	parallel group$1.ti,ab. (32580)

19	(crossover or cross over).ti,ab. (125730)

20	((assign$ or match or matched or allocation) adj5 (alternate or group$1 or intervention$1 or patient$1 or subject$1 or participant$1)).ti,ab. (420795)

21	(assigned or allocated).ti,ab. (497209)

22	(controlled adj7 (study or design or trial)).ti,ab. (456970)

23	(volunteer or volunteers).ti,ab. (284653)

24	human experiment/ (651172)

25	trial.ti. (408345)

26	or/7-25 (6411600)

27	(random$ adj sampl$ adj7 ("cross section$" or questionnaire$1 or survey$ or database$1)).ti,ab. not (comparative study/ or controlled study/ or randomi?ed controlled.ti,ab. or randomly assigned.ti,ab.) (9678)

28	Cross-sectional study/ not (randomized controlled trial/ or controlled clinical study/ or controlled study/ or randomi?ed controlled.ti,ab. or control group$1.ti,ab.) (368853)

29	(((case adj control$) and random$) not randomi?ed controlled).ti,ab. (21826)

30	(Systematic review not (trial or study)).ti. (267248)

31	(nonrandom$ not random$).ti,ab. (19102)

32	"Random field$".ti,ab. (2993)

33	(random cluster adj3 sampl$).ti,ab. (1604)

34	(review.ab. and review.pt.) not trial.ti. (1150148)

35	"we searched".ab. and (review.ti. or review.pt.) (50455)

36	"update review".ab. (137)

37	(databases adj4 searched).ab. (64166)

38	(rat or rats or mouse or mice or swine or porcine or murine or sheep or lambs or pigs or piglets or rabbit or rabbits or cat or cats or dog or dogs or cattle or bovine or monkey or monkeys or trout or marmoset$1).ti. and animal experiment/ (1232332)

39	Animal experiment/ not (human experiment/ or human/) (2587994)

40	or/27-39 (4401726)

41	26 not 40 (5654324)

42	6 and 41 (3855)

PsycInfo OVID

Database: APA PsycInfo <1806 to November Week 3 2023>

Search Strategy:

--------------------------------------------------------------------------------

1	post-covid-19 conditions/ (140)

2	(long* adj3 (covid or covid-19 or covid19 or sars cov 2)).mp. (476)

3	((covid* or "corona virus 2019" or "coronavirus2019" or SARS-CoV-2 or sars cov 2) adj3 (syndrome or persist* or lingering or chronic or ongoing or long-term or "long term" or long-haul or "long haul" or convalescen* or rehabilitat*)).ti. (190)

4	exp covid-19/ and sequela*.ti,ab. (276)

5	((Covid or Covid19 or "corona virus 2019" or "coronavirus 2019" or SARS-CoV-2 or "B.1.1.7" or "B.1.351" or "B.1.1.28" or "B.1.617" or "BA.1" or "BA.2" or "BA.3" or "BA.4" or "BA.5" or omicron or deltacron or "delta variant" or "delta subvariant" or "XBB.1.3") adj3 (PASC or sequela* or "post acute" or postacute or prolonged or "long haul*" or chronic or lingering or ongoing or persistent or "long term" or "more than 12 weeks" or "more than 24 weeks")).mp. [mp=title, abstract, heading word, table of contents, key concepts, original title, tests & measures, mesh word] (877)

6	or/1-5 (1399)

7	clinical trials/ (12263)

8	random:.tw. or placebo:.mp. or double-blind:.tw. (273444)

9	((treatment or control) adj3 group*).ab. (125944)

10	(allocat* adj5 group*).ab. (3242)

11	((clinical or control*) adj3 trial).ti,ab. (55610)

12	or/7-11 (371422)

13	6 and 12 (96)

AMED (OVID)

Database: AMED (Allied and Complementary Medicine) <1985 to October 2023>

Search Strategy:

--------------------------------------------------------------------------------

1	(long* adj3 (covid or covid-19 or covid19 or sars cov 2)).mp. (40)

2	((covid* or "corona virus 2019" or "coronavirus2019" or SARS-CoV-2 or sars cov 2) adj3 (syndrome or persist* or lingering or chronic or ongoing or long-term or "long term" or long-haul or "long haul" or convalescen* or rehabilitat*)).ti. (100)

3	((Covid or Covid19 or "corona virus 2019" or "coronavirus 2019" or SARS-CoV-2 or "B.1.1.7" or "B.1.351" or "B.1.1.28" or "B.1.617" or "BA.1" or "BA.2" or "BA.3" or "BA.4" or "BA.5" or omicron or deltacron or "delta variant" or "delta subvariant" or "XBB.1.3") adj3 (PASC or sequela* or "post acute" or postacute or prolonged or "long haul*" or chronic or lingering or ongoing or persistent or "long term" or "more than 12 weeks" or "more than 24 weeks")).mp. (80)

4	or/1-3 (159)

5	exp clinical trials/ (5249)

6	random:.tw. or placebo:.mp. or double-blind:.tw. (27785)

7	((treatment or control) adj3 group*).ab. (15197)

8	(allocat* adj5 group*).ab. (1155)

9	((clinical or control*) adj3 trial).ti,ab. (10341)

10	or/5-9 (37012)

11	4 and 10 (14)

CINAHL (EBSCO)

Thursday, November 30, 2023 8:45:56 PM

| **#** | **Query** | **Limiters/Expanders** | **Last Run Via** | **Results** |
| --- | --- | --- | --- | --- |
| S32 | S23 AND S31 | Search modes - Boolean/Phrase | Interface - EBSCOhost Research Databases Search Screen - Advanced Search Database - CINAHL | 733 |
| S31 | S24 OR S25 OR S26 OR S27 OR S30 | Search modes - Boolean/Phrase | Interface - EBSCOhost Research Databases Search Screen - Advanced Search Database - CINAHL | 7,822 |
| S30 | S28 AND S29 | Search modes - Boolean/Phrase | Interface - EBSCOhost Research Databases Search Screen - Advanced Search Database - CINAHL | 398 |
| S29 | TX sequela* | Search modes - Boolean/Phrase | Interface - EBSCOhost Research Databases Search Screen - Advanced Search Database - CINAHL | 17,889 |
| S28 | (MH "COVID-19+") | Search modes - Boolean/Phrase | Interface - EBSCOhost Research Databases Search Screen - Advanced Search Database - CINAHL | 45,583 |
| S27 | TX ((Covid or Covid19 or "corona virus 2019" or "coronavirus 2019" or SARS-CoV-2 or "B.1.1.7" or "B.1.351" or "B.1.1.28" or "B.1.617" or "BA.1" or "BA.2" or "BA.3" or "BA.4" or "BA.5" or omicron or deltacron or "delta variant" or "delta subvariant" or "XBB.1.3") N3 (PASC or sequela* or "post acute" or postacute or prolonged or "long haul*" or chronic or lingering or ongoing or persistent or "long term" or "more than 12 weeks" or "more than 24 weeks")) | Search modes - Boolean/Phrase | Interface - EBSCOhost Research Databases Search Screen - Advanced Search Database - CINAHL | 4,549 |
| S26 | TI ((covid* or "corona virus 2019" or "coronavirus2019" or SARS-CoV-2 or sars cov 2) N3 (syndrome or persist* or lingering or chronic or ongoing or long-term or "long term" or long-haul or "long haul" or convalescen* or rehabilitat*)) | Search modes - Boolean/Phrase | Interface - EBSCOhost Research Databases Search Screen - Advanced Search Database - CINAHL | 2,806 |
| S25 | TX (long* N3 (covid or covid-19 or covid19 or sars cov 2)) | Search modes - Boolean/Phrase | Interface - EBSCOhost Research Databases Search Screen - Advanced Search Database - CINAHL | 2,826 |
| S24 | (MH "Post-Acute COVID-19 Syndrome") | Search modes - Boolean/Phrase | Interface - EBSCOhost Research Databases Search Screen - Advanced Search Database - CINAHL | 1,047 |
| S23 | S22 NOT S21 | Expanders - Apply equivalent subjects Search modes - Boolean/Phrase | Interface - EBSCOhost Research Databases Search Screen - Advanced Search Database - CINAHL | 981,337 |
| S22 | S1 OR S2 OR S3 OR S4 OR S5 OR S6 OR S7 OR S8 OR S9 OR S10 OR S11 OR S12 OR S13 OR S14 OR S15 | Expanders - Apply equivalent subjects Search modes - Boolean/Phrase | Interface - EBSCOhost Research Databases Search Screen - Advanced Search Database - CINAHL | 1,029,252 |
| S21 | S19 NOT S20 | Expanders - Apply equivalent subjects Search modes - Boolean/Phrase | Interface - EBSCOhost Research Databases Search Screen - Advanced Search Database - CINAHL | 214,511 |
| S20 | MH (human) | Expanders - Apply equivalent subjects Search modes - Boolean/Phrase | Interface - EBSCOhost Research Databases Search Screen - Advanced Search Database - CINAHL | 2,730,925 |
| S19 | S16 OR S17 OR S18 | Expanders - Apply equivalent subjects Search modes - Boolean/Phrase | Interface - EBSCOhost Research Databases Search Screen - Advanced Search Database - CINAHL | 248,624 |
| S18 | TI (animal model*) | Expanders - Apply equivalent subjects Search modes - Boolean/Phrase | Interface - EBSCOhost Research Databases Search Screen - Advanced Search Database - CINAHL | 3,599 |
| S17 | MH (animal studies) | Expanders - Apply equivalent subjects Search modes - Boolean/Phrase | Interface - EBSCOhost Research Databases Search Screen - Advanced Search Database - CINAHL | 154,533 |
| S16 | MH animals+ | Expanders - Apply equivalent subjects Search modes - Boolean/Phrase | Interface - EBSCOhost Research Databases Search Screen - Advanced Search Database - CINAHL | 103,115 |
| S15 | AB (cluster W3 RCT) | Expanders - Apply equivalent subjects Search modes - Boolean/Phrase | Interface - EBSCOhost Research Databases Search Screen - Advanced Search Database - CINAHL | 503 |
| S14 | MH (crossover design) OR MH (comparative studies) | Expanders - Apply equivalent subjects Search modes - Boolean/Phrase | Interface - EBSCOhost Research Databases Search Screen - Advanced Search Database - CINAHL | 482,768 |
| S13 | AB (control W5 group) | Expanders - Apply equivalent subjects Search modes - Boolean/Phrase | Interface - EBSCOhost Research Databases Search Screen - Advanced Search Database - CINAHL | 146,556 |
| S12 | PT (randomized controlled trial) | Expanders - Apply equivalent subjects Search modes - Boolean/Phrase | Interface - EBSCOhost Research Databases Search Screen - Advanced Search Database - CINAHL | 153,885 |
| S11 | MH (placebos) | Expanders - Apply equivalent subjects Search modes - Boolean/Phrase | Interface - EBSCOhost Research Databases Search Screen - Advanced Search Database - CINAHL | 13,875 |
| S10 | MH (sample size) AND AB (assigned OR allocated OR control) | Expanders - Apply equivalent subjects Search modes - Boolean/Phrase | Interface - EBSCOhost Research Databases Search Screen - Advanced Search Database - CINAHL | 4,452 |
| S9 | TI (trial) | Expanders - Apply equivalent subjects Search modes - Boolean/Phrase | Interface - EBSCOhost Research Databases Search Screen - Advanced Search Database - CINAHL | 186,494 |
| S8 | AB (random*) | Expanders - Apply equivalent subjects Search modes - Boolean/Phrase | Interface - EBSCOhost Research Databases Search Screen - Advanced Search Database - CINAHL | 403,154 |
| S7 | TI (randomised OR randomized) | Expanders - Apply equivalent subjects Search modes - Boolean/Phrase | Interface - EBSCOhost Research Databases Search Screen - Advanced Search Database - CINAHL | 145,326 |
| S6 | MH cluster sample | Expanders - Apply equivalent subjects Search modes - Boolean/Phrase | Interface - EBSCOhost Research Databases Search Screen - Advanced Search Database - CINAHL | 5,356 |
| S5 | MH pretest-posttest design | Expanders - Apply equivalent subjects Search modes - Boolean/Phrase | Interface - EBSCOhost Research Databases Search Screen - Advanced Search Database - CINAHL | 54,200 |
| S4 | MH random assignment | Expanders - Apply equivalent subjects Search modes - Boolean/Phrase | Interface - EBSCOhost Research Databases Search Screen - Advanced Search Database - CINAHL | 82,143 |
| S3 | MH single-blind studies | Expanders - Apply equivalent subjects Search modes - Boolean/Phrase | Interface - EBSCOhost Research Databases Search Screen - Advanced Search Database - CINAHL | 16,069 |
| S2 | MH double-blind studies | Expanders - Apply equivalent subjects Search modes - Boolean/Phrase | Interface - EBSCOhost Research Databases Search Screen - Advanced Search Database - CINAHL | 54,318 |
| S1 | MH randomized controlled trials | Expanders - Apply equivalent subjects Search modes - Boolean/Phrase | Interface - EBSCOhost Research Databases Search Screen - Advanced Search Database - CINAHL | 140,118 |

Cochrane Library (Wiley)

Search Name:	2023-11-30 Long Covid revised

Date Run:	30/11/2023 22:23:02

Comment:

ID	Search	Hits

#1	MeSH descriptor: [Post-Acute COVID-19 Syndrome] explode all trees	78

#2	(((covid* or "corona virus 2019" or "coronavirus2019" or SARS-CoV-2 or sars cov 2) NEAR/3 (syndrome or persist* or lingering or chronic or ongoing or long-term or "long term" or long-haul or "long haul" or convalescen* or rehabilitat*))):ti (Word variations have been searched)	1294

#3	((Covid or Covid19 or "corona virus 2019" or "coronavirus 2019" or SARS-CoV-2 or "B.1.1.7" or "B.1.351" or "B.1.1.28" or "B.1.617" or "BA.1" or "BA.2" or "BA.3" or "BA.4" or "BA.5" or omicron or deltacron or "delta variant" or "delta subvariant" or "XBB.1.3") NEAR/3 (PASC or sequela* or "post acute" or postacute or prolonged or "long haul" or chronic or lingering or ongoing or persistent or "long term" or "more than 12 weeks" or "more than 24 weeks"))	608

#4	MeSH descriptor: [COVID-19] explode all trees	4984

#5	MeSH descriptor: [SARS-CoV-2] explode all trees	2457

#6	#4 or #5	5198

#7	sequela*	5641

#8	#6 and #7	94

#9	long covid	1900

#10	long NEAR/3 (covid or covid19 or covid 19 or sars cov 2)	3808

#11	(long-haul or long-term) NEAR/3 (covid or covid19 or covid 19 or sars cov 2)	2171

#12	#1 or #2 or #3 or #8 or #9 or #10 or #11 in Trials	6088
